# Cost-Effectiveness of Daraxonrasib vs Chemotherapy in Previously Treated Metastatic Pancreatic Adenocarcinoma

**DOI:** 10.64898/2026.09.16.26363246

**Authors:** Sunwoo Han, Natallia Starasvetskaya, Gretel Terrero, Peter J. Hosein, Gilberto de Lima Lopes

**Author notes:** Authors contributed equally as first authors (Sunwoo Han, Natallia Starasvetskaya). Authors contributed equally as senior authors (Peter J. Hosein, Gilberto de Lima Lopes).

## Abstract

**Introduction:** Daraxonrasib is the first broad RAS-targeted medicine approved for metastatic pancreatic adenocarcinoma. In RASolute 302, it approximately doubled median overall survival in the prespecified RAS G12 population. The US wholesale acquisition cost (WAC) is $39,800 per 30-day supply. Contemporary oncology launch prices have risen substantially, and many recently launched drugs have been priced above value-based benchmarks. The value of a genuinely transformative oncology therapy at this price is uncertain.

**Objective:** To estimate the cost-effectiveness of daraxonrasib compared with investigator’s-choice chemotherapy in previously treated metastatic pancreatic adenocarcinoma using publicly available clinical and economic data.

**Design, Setting, and Participants:** Economic evaluation using a 3-state partitioned survival model informed by the international randomized phase 3 RASolute 302 trial. Published Kaplan-Meier curves and numbers at risk were used to reconstruct pseudo-individual patient data. The RAS G12 population (n=459), corresponding to the trial’s dual primary efficacy end points, was the primary economic population; the overall randomized population (n=500) was analyzed in parallel. Analyses were conducted in 2026 from a US health sector perspective.

Exposures: Daraxonrasib 300 mg orally once daily versus investigator’s-choice chemotherapy, weighted to the observed trial regimen distribution.

**Main Outcomes and Measures:** Incremental life-years, quality-adjusted life-years (QALYs), costs, incremental cost-effectiveness ratio (ICER), incremental net monetary benefit, and the 30-day daraxonrasib price consistent with willingness-to-pay thresholds of $100,000, $150,000, $200,000, and $255,000/QALY. Costs were expressed in 2026 US dollars and discounted at 3% annually.

**Results:** In the RAS G12 population, daraxonrasib generated 0.652 additional life-years and 0.506 additional QALYs at an incremental cost of $253,171, yielding an ICER of $500,637/QALY. The corresponding overall-population ICER was $542,255/QALY. A 10-year horizon changed the RAS G12 ICER to $497,270/QALY; using curve-specific distributions selected by Akaike information criterion produced $493,895/QALY. In 5000 probabilistic simulations, the probability of cost-effectiveness was 0% at $150,000 and $200,000/QALY and 0.1% at $255,000/QALY. Threshold prices in the RAS G12 population were $14,060 per 30 days at $150,000/QALY and $17,730 at $200,000/QALY, reductions of 64.7% and 55.5% from WAC. An independent secondary implementation using separately digitized survival curves and piecewise-exponential extrapolation produced a higher ICER; after calibrating drug exposure to observed treatment duration, the estimate remained approximately $676,000/QALY, with most of the difference attributable to the extrapolated treatment-exposure tail.

**Conclusions and Relevance:** Daraxonrasib nearly doubled median overall survival in previously treated metastatic pancreatic adenocarcinoma, in which median survival with standard chemotherapy was about 7 months, and should remain available to eligible patients. At WAC, daraxonrasib costs approximately $500,000 per QALY, well above conventional US benchmarks. Because actual net acquisition prices may reflect confidential discounts or rebates, the ICER based on WAC may exceed the ICER at the net price. Threshold list prices consistent with $150,000-$200,000/QALY were $14,060-$17,730 per 30 days (a 55%-65% reduction from WAC), offered as reference points for pricing and coverage discussions rather than recommended prices.

## Introduction

Metastatic pancreatic ductal adenocarcinoma remains one of the most lethal solid tumors, and outcomes after progression on first-line chemotherapy have historically been poor. More than 90% of pancreatic ductal adenocarcinomas have oncogenic RAS alterations, yet direct pharmacologic inhibition of RAS remained elusive for decades.^1^

RASolute 302 changed that clinical landscape. In the prespecified RAS G12 population, median overall survival was 13.2 months with daraxonrasib and 6.6 months with investigator’s choice chemotherapy (hazard ratio, 0.40; 95% CI, 0.30-0.54); median progression-free survival was 7.3 vs 3.5 months. The benefit was similar in the overall randomized population, and patient-reported pain and global health status deteriorated later with daraxonrasib.^1^

The US Food and Drug Administration approved daraxonrasib on August 26, 2026, for adults with metastatic pancreatic adenocarcinoma who had received at least 1 prior systemic therapy or were not candidates for multiagent systemic therapy. The manufacturer reported a wholesale acquisition cost (WAC) of $39,800 per 30-day supply, or $477,600 annualized.^2,3^

We evaluated whether that magnitude of clinical benefit justifies the launch price and estimated the price at which daraxonrasib would meet commonly used US cost-effectiveness thresholds ($100,000-$200,000/QALY, consistent with contemporary US value-assessment frameworks). We designed the model as an independent, fully auditable analysis using public data.^4^

## Methods

### Model Overview

We constructed a 3-state partitioned survival model with progression-free, progressed, and dead states, weekly cycles, trapezoidal half-cycle correction, and a 5-year reference horizon. A 10-year horizon was examined as a scenario and produced materially identical results, supporting the 5-year horizon as an effective lifetime horizon in this population. The perspective was the US health sector. Costs and health outcomes were discounted 3% annually. Reporting follows CHEERS 2022.^5^

Institutional review board approval was not required because the analysis used published aggregate trial data and reconstructed pseudo-individual survival data without access to identifiable participants. Model code, reconstruction outputs, and the frozen parameter register are available from the corresponding author upon reasonable request.

### Population and Comparators

The primary economic population was the RAS G12 population (n=459), corresponding to the population in which the trial’s dual primary end points of overall and progression-free survival were prespecified. A parallel analysis used all 500 randomized patients because the approved indication is not restricted by RAS genotype. Investigator’s-choice chemotherapy was weighted to the observed trial distribution: gemcitabine plus nab-paclitaxel (56.5%), liposomal irinotecan plus fluorouracil and leucovorin (32.7%), modified FOLFIRINOX (5.6%), and FOLFOX (5.1%).^1^

### Survival Reconstruction and Extrapolation

Published Kaplan-Meier curves for overall and progression-free survival in each arm and population were extracted from the vector graphics of the NEJM report. Pseudo-individual patient data were reconstructed using the Guyot method, constrained by published numbers at risk and total event counts.^6^

Reconstruction validity was assessed by comparing reconstructed sample sizes, event totals, medians, landmark survival, and the published Kaplan-Meier curves. All 8 reconstructed curves exactly matched sample size, event totals, and reported numbers at risk; curve root-mean-square error was less than 0.9 percentage points. Weibull, exponential, log-normal, log-logistic, and Gompertz models were fitted to each reconstructed curve. Weibull was prespecified for the reference case to maintain a common parametric family across arms and end points; curve-specific models selected by Akaike information criterion were examined in sensitivity analysis. The latter selected Weibull for daraxonrasib OS and PFS and chemotherapy OS, and log-normal for chemotherapy PFS in the RAS G12 population.

### Treatment Exposure, Costs, and Utilities

Treatment exposure was calibrated to the reported median treatment duration of 6.2 months with daraxonrasib and regimen-specific medians of 1.5 to 3.2 months with chemotherapy, scaled by the proportions receiving at least 1 dose (97.2% and 84.9%). Because 42.3% of daraxonrasib-treated patients remained on therapy at the data cutoff, this fixed-median approach is a conservative simplification rather than a fitted time-on-treatment distribution. No additional final oral fill was charged at discontinuation; this reference assumption avoids attributing an unsupported full or partial prescription wastage cost.^1^

Daraxonrasib was costed at WAC ($39,800/30 days). Chemotherapy acquisition costs were calculated bottom-up from trial dosing and the realized regimen distribution using July 2026

Medicare Part B average sales price payment limits, vial sizes, and billed vial wastage; administration costs used 2026 Medicare Physician Fee Schedule national nonfacility rates. The resulting mix-weighted comparator cost was $9,655 for drugs plus $847 for administration per 30 days.^7^

Routine follow-up costs were $550/month while progression-free and $1,128/month after progression, based on a US public-payer pancreatic cancer economic evaluation and inflated to 2026 dollars. Terminal care was $20,033 as a one-time cost. Subsequent-therapy costs were omitted from the reference case because RASolute 302 did not report arm-specific post-progression treatment use, avoiding unsupported differential costs and possible double counting.^8,9^

Grade 3 or higher treatment-related adverse events were observed in 43.6% of daraxonrasib-treated patients and 57.5% of chemotherapy-treated patients. Trial event-specific rates were combined with Medicare claims estimates of excess costs associated with hematologic and nonhematologic adverse events, yielding expected adverse-event costs of $2,343 and $4,758 per patient, respectively.^1,10^

Because RASolute 302 collected EORTC QLQ-C30 and QLQ-PAN26 rather than a preference-based utility instrument, utilities were borrowed from pancreatic cancer economic literature: 0.85 while progression-free and 0.73 after progression, applied identically to both treatment arms.^9^ These relatively high progression-free utilities favor daraxonrasib because it generates more progression-free survival. A +0.05 treatment-specific progression-free utility increment for daraxonrasib was examined as a favorable scenario, supported qualitatively by the trial’s patient-reported outcome advantage.^1^ Because this borrowed, arm-identical approach does not capture the trial’s own patient-reported outcome advantage for daraxonrasib, it is more likely to understate than overstate the drug’s true QALY benefit.

### Analyses

The primary outcome was incremental cost per QALY. We also calculated incremental life-years, incremental net monetary benefit, and threshold prices at $100,000, $150,000, $200,000, and $255,000/QALY. The $100,000-$150,000 range corresponds to commonly used US health-benefit price benchmarks, with $200,000 included as a more permissive upper scenario.11 The $255,000/QALY value is included as an exploratory severity-adjusted benchmark rather than a conventional US threshold. Medicare has no statutory or regulatory cost-per-QALY threshold: the Affordable Care Act (Section 1182) and subsequent Inflation Reduction Act guidance explicitly bar the Centers for Medicare & Medicaid Services from using QALY-based cost-effectiveness thresholds in coverage determinations or in setting a negotiated Maximum Fair Price. The thresholds used here are drawn from non-governmental US value-assessment conventions (e.g., the Institute for Clinical and Economic Review) and should be read as reference benchmarks, not as a description of Medicare policy or an implied negotiation target.

One-way sensitivity analysis varied key inputs over prespecified plausible ranges. Probabilistic sensitivity analysis used 5000 Monte Carlo simulations, sampling uncertainty in reconstructed survival parameters, utilities, treatment duration, and cost inputs. A sequential stress test cumulatively applied assumptions favorable to daraxonrasib, including a 35% net-price discount, alternative survival extrapolation, 10-year horizon, treatment-specific progression-free utility advantage, higher comparator acquisition cost, and a societal cost assigned to infusion time and travel.

As an independent structural validation, one of the authors independently digitized the published RAS G12 and overall-population Kaplan-Meier curves and implemented a previously used cost-effectiveness framework without access to the reference model outputs. That implementation selected piecewise-exponential survival extrapolation based on goodness-of-fit and visual assessment. The initial implementation costed treatment through progression; for model reconciliation, treatment exposure was recalibrated separately to the observed median treatment durations while retaining the independently reconstructed survival curves and other model assumptions.

## Results

### Survival Reconstruction

The reconstructed RAS G12 curves reproduced the published medians closely: 13.25 vs 13.2 months for daraxonrasib OS, 6.72 vs 6.6 months for chemotherapy OS, 7.31 vs 7.3 months for daraxonrasib PFS, and 3.49 vs 3.5 months for chemotherapy PFS. At 5 years, the reference Weibull model generated 0.652 discounted incremental life-years for daraxonrasib. The curve-specific AIC strategy yielded the same incremental life-years and only modest differences in QALYs and costs.

### Reference-Case Cost-Effectiveness

In the RAS G12 population, daraxonrasib generated 1.037 QALYs compared with 0.532 for chemotherapy, for an incremental 0.506 QALYs. Mean modeled costs were $308,785 and $55,615, respectively, producing an incremental cost of $253,171 and an ICER of $500,637/QALY. In the overall randomized population, the corresponding values were 0.472 incremental QALYs, $255,881 incremental cost, and $542,255/QALY.

The 10-year horizon produced an ICER of $497,270/QALY in the RAS G12 population. Allowing each RAS G12 curve to use its AIC-preferred parametric family produced $493,895/QALY. Thus, neither the reference horizon nor reasonable parametric model selection materially altered the result.

### Independent Model Validation

The independently implemented analysis produced 0.47 incremental QALYs and approximately 0.64 incremental life-years in the RAS G12 population, closely matching the reference model estimates of 0.506 QALYs and 0.652 life-years. When drug acquisition was costed through progression, its ICER was $771,670/QALY. Separating treatment exposure from PFS and calibrating exposure to the observed treatment-duration medians reduced the estimate to approximately $675,700/QALY. A stepwise reconciliation showed that most of the remaining difference from the reference estimate was explained by the longer mean treatment exposure implied by the piecewise-exponential tail; harmonizing expected treatment exposure brought the independent framework to approximately $526,000/QALY before smaller differences in QALY and non-drug costs.

### Sensitivity and Probabilistic Analyses

The largest one-way driver was daraxonrasib time on treatment (ICER range, $396,757-$612,612/QALY), followed by daraxonrasib median OS ($431,796-$610,034), drug price ($337,989-$500,637), progressed-state utility ($448,117-$567,103), and chemotherapy median OS ($449,400-$540,332). Terminal-care costs had minimal influence.

In probabilistic analysis, the probability that daraxonrasib was cost-effective was 0% at $150,000/QALY, 0% at $200,000/QALY, and 0.12% at $255,000/QALY. The probability approached 50% only around $500,000/QALY. Incremental net monetary benefit remained negative at all thresholds through $255,000/QALY.

### Threshold Price and Assumption Stress Test

The 30-day daraxonrasib price compatible with $150,000/QALY was $14,060, a 64.7% reduction from launch WAC. At $200,000/QALY, the threshold price was $17,730, a 55.5% reduction. Corresponding overall-population prices were $13,243 and $16,628.

When favorable assumptions were stacked sequentially, the ICER declined from $500,637/QALY in the reference case to $310,881 after applying a 35% net-price discount, $271,814 with log-normal extrapolation, and $209,443 with a 10-year horizon. Adding a daraxonrasib-specific progression-free utility advantage, a higher comparator drug cost, and societal infusion/travel costs produced a fully stressed ICER of $191,129/QALY. This scenario is not a reference estimate; rather, it identifies how many simultaneously favorable assumptions are required before the ratio approaches $200,000/QALY.

## Discussion

Daraxonrasib presents an unusual test of value in oncology. In RASolute 302, it approximately doubled median overall survival in previously treated metastatic pancreatic cancer, a magnitude of benefit that is uncommon in this disease and is clearly clinically relevant. Yet at $39,800 per 30-day supply, the reference-case ICER remains approximately $500,000/QALY. (An ICER expresses the extra cost required to gain one quality-adjusted life-year relative to the comparator; it is a tool for comparing value across treatments and informing price negotiation, not a statement about the value of an individual patient’s life.) This is not a conclusion driven by a small clinical effect: the economic tension persists despite a 6.6-month difference in median survival and approximately 0.65 modeled life-years gained. Daraxonrasib was also better tolerated than chemotherapy, with fewer grade ≥3 treatment-related adverse events (43.6% vs 57.5%)1 — a clinically meaningful advantage, independent of cost, for patients who have already progressed on first-line therapy.

Daraxonrasib’s annualized WAC is high but lies within the range of contemporary oncology launch prices. Inflation-adjusted net launch prices for new cancer drugs rose from approximately $100,000 to approximately $200,000 per year of treatment between 2008 and 2022, with only a weak and inconsistent relationship to the magnitude of clinical benefit across that period.^17^ For context, ICER’s 2025 Launch Price and Access Report found median oncology launch list prices of $357,623 in 2022, $346,980 in 2023, and $442,645 in 2024, with corresponding estimated median net prices of $316,515, $302,381, and $403,823. Among 23 recently launched drugs that had previously undergone ICER review, 16 had estimated net prices above the upper bound of ICER’s health-benefit price benchmark.^18^ Daraxonrasib’s annualized WAC of $477,600 is therefore above the recent median oncology list price but within the same order of magnitude. This context does not change the ICER we report; rather, it places daraxonrasib within a broader market pattern in which launch prices can exceed value-based benchmarks.

The most decision-relevant finding is therefore not the precise ICER but the threshold price. A price near $14,000-$18,000 per 30 days would align daraxonrasib with thresholds of $150,000-$200,000/QALY, implying a reduction of approximately 55% to 65% from WAC. This threshold is a benchmark for value at list price, not a recommended price cut: manufacturers typically negotiate confidential rebates with payers and pharmacy benefit managers, so daraxonrasib’s actual net price today is unknown and may already be materially below WAC. Threshold pricing is also less sensitive than the headline ICER to uncertain downstream costs, utility assumptions, and survival extrapolation and translates directly into a quantity relevant to payers and manufacturers.

Several features of our analyses intentionally bias the reference case toward daraxonrasib. The progression-free utility of 0.85 is generous for metastatic pancreatic cancer and gives substantial quality-adjusted value to the additional progression-free survival produced by the drug. The comparator reflects the actual trial mix rather than the least expensive chemotherapy option. No unsupported subsequent therapy penalty is assigned to daraxonrasib. We also removed an assumed unused final oral fill because actual refill timing at discontinuation is unknown. Despite these choices, the ICER remains far above commonly used US benchmarks.

The analysis also clarifies which information would most reduce uncertainty. Time on treatment with daraxonrasib is the largest deterministic driver because acquisition cost accumulates while therapy continues. RASolute 302 reported a median treatment duration but not a treatment-duration Kaplan-Meier curve; 42.3% of daraxonrasib-treated patients were still receiving therapy at the data cutoff. Publication of mature treatment-exposure data may therefore be more economically informative than additional precision around routine care or terminal costs, which have little effect on the result.

The independent implementation strengthens this interpretation. Despite different curve extraction, extrapolation, and model architecture, the estimated incremental health benefit was very similar.

The higher independent ICER was driven primarily by the mean treatment duration generated by the extrapolated treatment-exposure tail. This divergence is decision-relevant rather than a failure of replication: it identifies mature time-on-treatment data as the principal unresolved structural uncertainty and shows that plausible alternative implementations remain far above conventional US willingness-to-pay thresholds.

The sequential stress test has a different purpose from ordinary sensitivity analysis. Cost-effectiveness models permit many individually defensible choices, and industry sponsorship has been associated with ICERs approximately one-third lower than non-sponsored analyses.^4^

Rather than implying that any single alternative assumption is biased, the stress test quantifies the cumulative analytic latitude available. In this case, daraxonrasib approaches $200,000/QALY only after multiple assumptions favorable to the intervention are applied simultaneously, including a large net-price discount. That finding reinforces the importance of transparent parameter provenance and public code. For a payer or policy audience, per-patient value (ICER) is only half the picture. Pancreatic cancer is not rare (approximately 66,000 new US diagnoses annually), so even a well-characterized per-patient ICER leaves open the aggregate budget impact that a Medicare or commercial payer would actually weigh in coverage and utilization-management decisions.

### Limitations

Like all health-economic assessments, our study has limitations. First, survival follow-up in RASolute 302 remains immature, so long-term benefit requires extrapolation. We mitigated this by reconstructing the published Kaplan-Meier curves, testing multiple parametric families, and showing little difference between the 5– and 10-year RAS G12 results. Second, pseudo-individual patient data reconstructed from published curves cannot reproduce the trial’s patient-level stratification factors; reconstructed unstratified hazard ratios are therefore validation metrics rather than substitutes for the published stratified estimates. Third, utilities were borrowed because the trial did not report a preference-based instrument. Fourth, WAC is not net price; confidential rebates would reduce the true acquisition cost, which is precisely why threshold prices and a discount scenario are reported. Fifth, treatment-duration and chemotherapy regimen-distribution inputs were available from the overall safety population and were applied to the RAS G12 economic analysis.

Sixth, arm-specific post-progression therapy was not reported and was omitted from the reference case. Seventh, the independent validation model was available as a separate implementation rather than a fully harmonized executable rerun; its role is therefore structural validation rather than pooled estimation. Finally, the approved indication includes patients who are not candidates for multiagent systemic therapy without prior treatment; no randomized evidence supports extrapolating this analysis to that population. These patients are clinically distinct (typically older, poorer performance status, ineligible for the trial’s chemotherapy regimens), and their appropriate comparator may be best supportive care or single-agent gemcitabine rather than the trial’s chemotherapy mix — which could make the ICER either more or less favorable in that subgroup, in either direction, and readers should not assume it generalizes.

Most importantly, a single willingness-to-pay threshold may fit this population poorly. Patients in RASolute 302 had a median overall survival of 6.6 months with standard chemotherapy and limited proven options after prior systemic therapy. The International Society for Pharmacoeconomics and Outcomes Research Special Task Force identified severity of disease, the value of hope, and real option value as elements that conventional cost-effectiveness may omit^13^, and the generalized risk-adjusted cost-effectiveness framework suggests that risk-averse individuals may value health gains differently when the baseline state is severe and near-terminal.^14,15^ Revolution Medicines reported company-wide research and development expenses of $592.2 million in 2024 and $987.3 million in 2025 and an accumulated deficit of $2.9 billion at the end of 2025, across a portfolio in which daraxonrasib is the lead asset.^16^ These company-wide figures cannot be attributed to daraxonrasib and are not inputs to this cost-effectiveness analysis; they illustrate the broader financing context for oncology innovation. A health system that rewards genuinely large clinical gains while maintaining affordability is a legitimate social goal.

## Conclusions

Daraxonrasib nearly doubles overall survival in one of the deadliest cancers and should be available to every eligible patient. The United States has no universally accepted cost-effectiveness threshold. Values of $100,000 to $150,000 per QALY are conventional but not authoritative, and severity-adjusted frameworks can support higher willingness to pay for a population with a standard-treatment median survival of about seven months. At launch WAC, daraxonrasib costs approximately $500,000/QALY. Threshold prices in the RAS G12 population were $14,060 per 30 days at $150,000/QALY and $17,730 at $200,000/QALY, reductions of 64.7% and 55.5% from WAC. These estimates are offered as an independent reference point for the pricing, coverage, and negotiation decisions that will determine how widely that benefit is delivered. They are not a recommendation to restrict access, and they should not be read as a description of what Medicare is permitted to do under current law.

## Tables

**Table 1.** Reference-Case Model Inputs.

| Parameter | Reference value | Source/rationale |
| --- | --- | --- |
| Clinical efficacy, RAS G12 | OS 13.2 vs 6.6 mo; PFS 7.3 vs 3.5 mo | RASolute 302 |
| Daraxonrasib WAC | \$39,800 per 30 d | Manufacturer / SEC filing |
| Comparator drugs | \$9,655 per 30 d | CMS July 2026 ASP; trial mix |
| Comparator administration | \$847 per 30 d | CY 2026 MPFS |
| Median treatment duration | 6.2 mo daraxonrasib; 1.5-3.2 mo chemotherapy | RASolute 302 |
| Utility, progression-free | 0.85 | Published pancreatic cancer CEA |
| Utility, progressed | 0.73 | Published pancreatic cancer CEA |
| PF follow-up | \$550/mo | Published US public-payer CEA; 2026-adjusted |
| Progressed follow-up | \$1,128/mo | Published US public-payer CEA; 2026-adjusted |
| Terminal care | \$20,033 one time | Published pancreatic cancer CEA; 2026-adjusted |
| Expected grade $\geq 3$ TRAE cost | \$2,343 dara; \$4,758 chemotherapy | RASolute 302 + Medicare claims |
| Subsequent therapy | \$0 differential in reference case | Not reported by trial; omitted to avoid unsupported differential |
| Unused final oral fill | 0 additional fills | Frozen structural assumption |
| Discount rate | 3% annually | Reference case |

**Table 2.** Base-Case and Survival Scenario Results.

| **Population / scenario** | **Incremental QALYs** | **Incremental cost, \$** | **ICER, \$/QALY** |
| --- | --- | --- | --- |
| G12, Weibull, 5 y | 0.506 | 253,171 | 500,637 |
| G12, Weibull, 10 y | 0.510 | 253,377 | 497,270 |
| G12, AIC-preferred, 5 y | 0.502 | 247,715 | 493,895 |
| G12, AIC-preferred, 10 y | 0.505 | 247,922 | 490,552 |
| Overall, Weibull, 5 y | 0.472 | 255,881 | 542,255 |

**Table 3.** Threshold Daraxonrasib Price.

| **Population** | **WTP, \$/QALY** | **Price per 30 d, \$** | **Reduction from WAC, %** |
| --- | --- | --- | --- |
| G12 | 100,000 | 10,389 | 73.9 |
| G12 | 150,000 | 14,060 | 64.7 |
| G12 | 200,000 | 17,730 | 55.5 |
| G12 | 255,000 | 21,768 | 45.3 |
| Overall | 100,000 | 9,858 | 75.2 |
| Overall | 150,000 | 13,243 | 66.7 |
| Overall | 200,000 | 16,628 | 58.2 |
| Overall | 255,000 | 20,352 | 48.9 |

## Figure Legends

**Figure 1.**
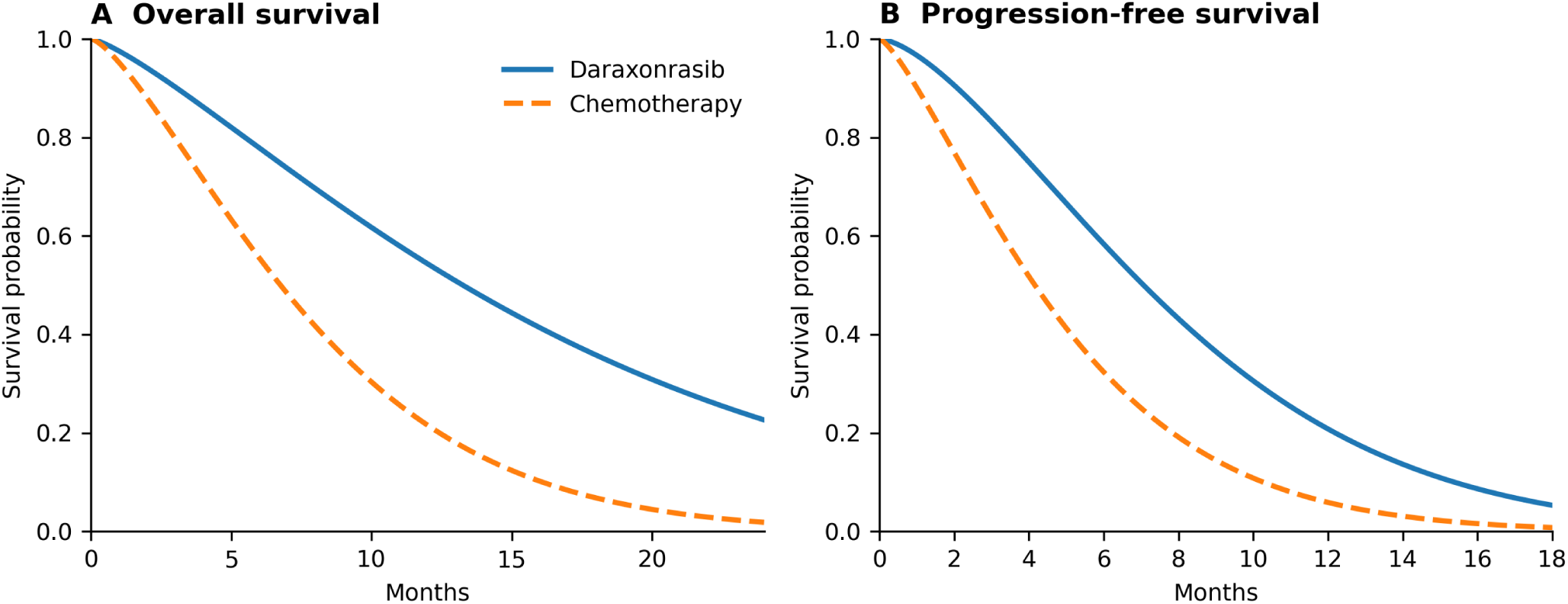
Parametric survival curves in the RAS G12 population. Reference-case Weibull models fitted to pseudo-individual patient data reconstructed from the published RASolute 302 Kaplan-Meier curves and numbers at risk.

**Figure 2.**
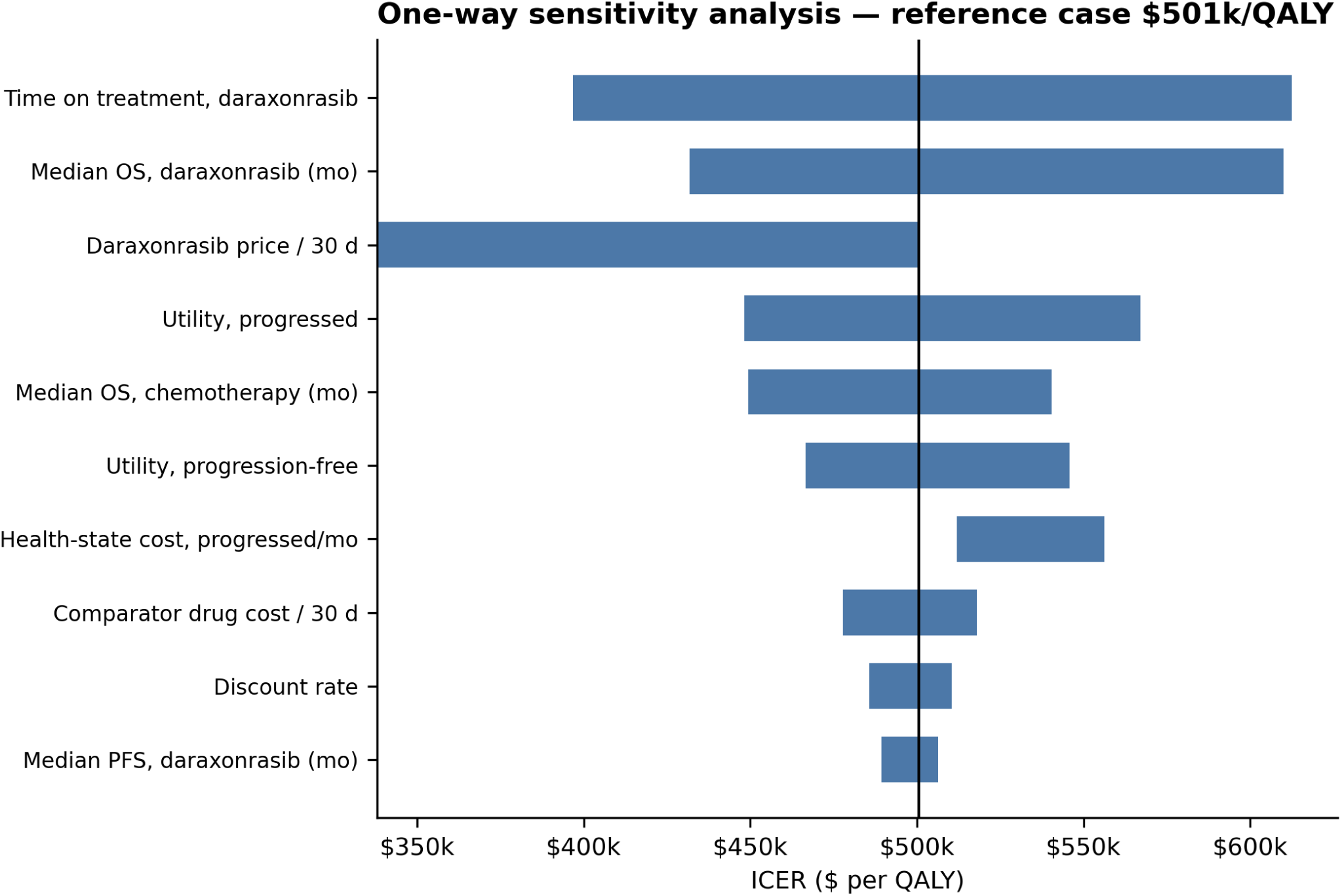
One-way deterministic sensitivity analysis in the RAS G12 population. The vertical line indicates the reference-case ICER of $500,637/QALY. The 10 parameters with the largest ICER ranges are shown.

**Figure 3.**
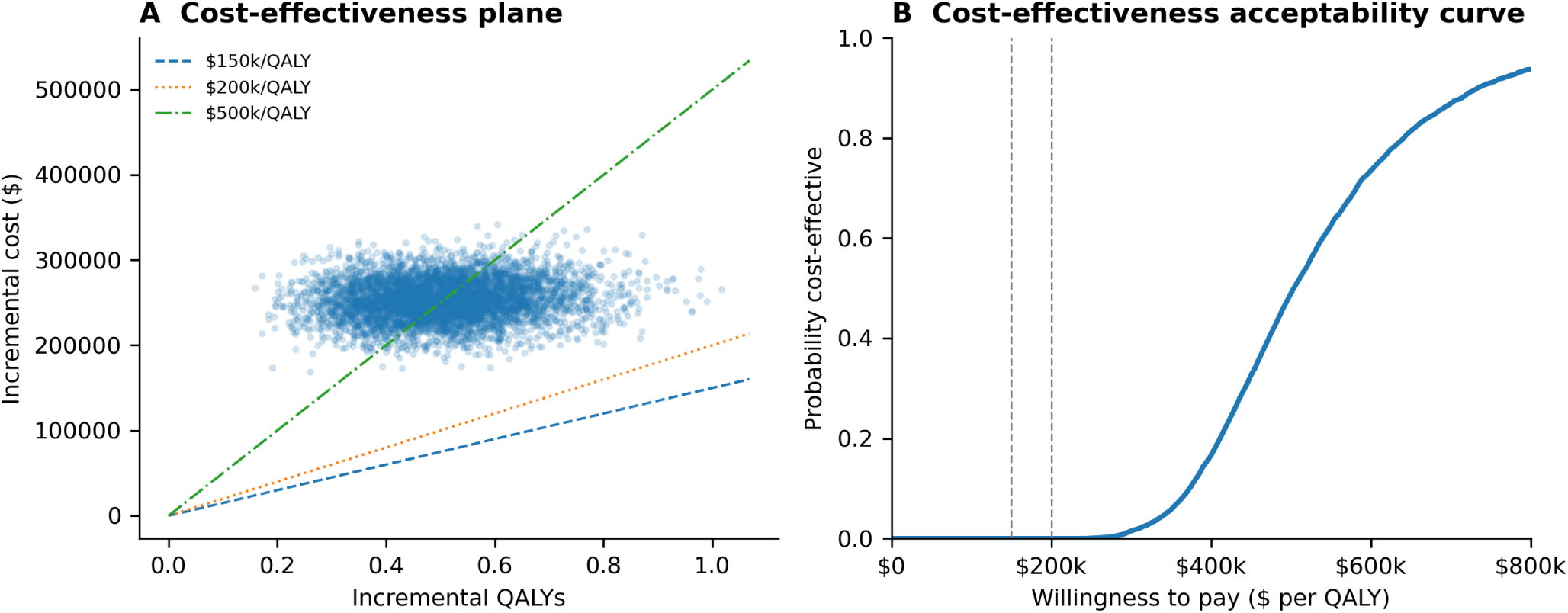
Probabilistic sensitivity analysis, RAS G12 population (5000 iterations). A, Cost-effectiveness plane. B, Cost-effectiveness acceptability curve.

**Figure 4.**
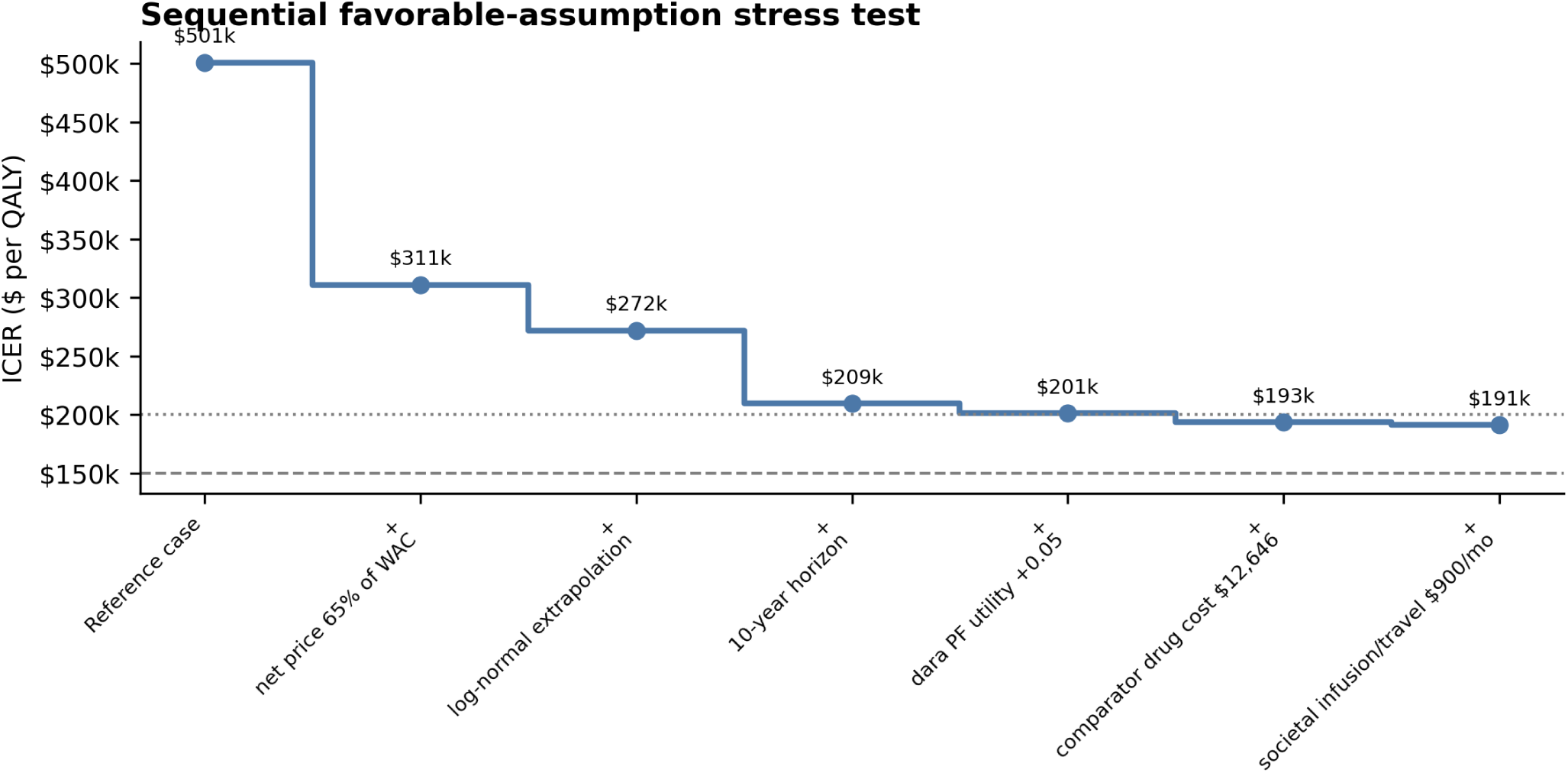
Sequential favorable-assumption stress test. Assumptions are cumulatively added to the reference model to quantify the analytic conditions required for the ICER to approach conventional upper willingness-to-pay thresholds. The fully stressed scenario is not a reference estimate.

## Data Availability

All data produced in the present study are available upon reasonable request to the authors

## Article Information

Data Sharing Statement: The reconstructed pseudo-individual survival data, frozen parameter register, analytic code, model outputs, and independent model reconciliation materials are included in the preprint reproducibility package and are intended for public deposition concurrently with the preprint.

## Notes

### Competing Interest Statement

Authors have participated in clinical trials run by revolutions medicine, no personal payments.

